# Quantifying structural connectivity in brain tumor patients

**DOI:** 10.1101/2021.03.19.21253837

**Authors:** Yiran Wei, Chao Li, Stephen John Price

**Affiliations:** Department of Clinical Neurosciences, University of Cambridge; Department of Neurosurgery, Shanghai General Hospital, Shanghai Jiao Tong University School of Medicine

**Author notes:** { }. Accepted by MICCAI 2021.

**Keywords:** brain networks, structural connectivity, brain tumor, tractography, diffusion MRI

## Abstract

Brain tumors are characterised by infiltration along the white matter tracts, posing significant challenges to precise treatment. Mounting evidence shows that an infiltrative tumor can interfere with the brain network diffusely. Therefore, quantifying structural connectivity has potential to identify tumor invasion and stratify patients more accurately. The tract-based statistics (TBSS) is widely used to measure the white matter integrity. This voxel-wise method, however, cannot directly quantify the connectivity of brain regions. Tractography is a fiber tracking approach, which has been widely used to quantify brain connectivity. However, the performance of tractography on the brain with tumors is biased by the tumor mass effect. A robust method of quantifying the structural connectivity in brain tumor patients is still lacking. Here we propose a method which could provide robust estimation of tract strength for brain tumor patients. Specifically, we firstly construct an unbiased tract template in healthy subjects using tractography. The voxel projection procedure of TBSS is employed to quantify the tract connectivity in patients, based on the location of each tract fiber from the template. To further improve the standard TBSS, we propose an approach of iterative projection of tract voxels, under the guidance of tract orientation measured by voxel-wise eigenvectors. Compared to the conventional tractography methods, our approach is more sensitive in reflecting functional relevance. Further, the different extent of network disruption revealed by our approach correspond to the clinical prior knowledge of tumor histology. The proposed method could provide a robust estimation of the structural connectivity for brain tumor patients.

## 1 Introduction

### 1.1 Brain tumors and white matter tracts

A brain tumor refers to a mass lesion identified within the brain or related structures. Among them, gliomas and meningiomas are the most common primary tumor types in adults. Impacts of tumors are characterized by diffuse infiltration and interference with the white matter tracts [24, 26], the neuronal fiber bundles forming a complex network connecting cortical regions. As a result, the tumor infiltration along white matter tracts may lead to structural disturbance of the brain[13].

Brain tumor patients frequently demonstrate neurological deficits that are not directly explained by the focal lesion [8]. Therefore, it is increasingly accepted that brain tumors may cause broader impacts to the global brain beyond the focal site through the white matter tracts [1]. Therefore, accurately measuring structural connectivity strength offers a promising imaging surrogate to detect the tumor-related brain alterations. Further, the pre-treatment mapping of the structural connectivity provides significance for the planning of both surgery and radiotherapy[18]

### 1.2 Connectivity strength measurement

Recent development of neuroimaging techniques have facilitated characterization of brain connectivity at the whole-brain level, based on diffusion MRI (dMRI). The derived fractional anisotropy (FA) map is commonly used to measure the tract strength. The tract-based spatial statistics (TBSS) is a method to estimate the strength of specific tracts using a FA skeleton derived by mapping the local maxima FA voxels to the template [19]. As a voxel-based method, however, TBSS fails to consider the spatial continuity of the fiber pathway, which has difficulties in multiple testing and tracking the fibers that tumor infiltrates along [4]. Further, as the projection procedure in TBSS is purely based on the FA values, the traditional TBSS does not consider the orientation of the tract fibers that are frequently affected in brain tumor patients.

Tractography is a widely-used fiber tracking method to measure the tract connectivity. This method has the advantage of detecting the fiber pathway with spatial consistency. Performing tractography on brain tumor patients, however, has the below challenges: 1) The tract pathways in vicinity to the tumor are often anatomically deviated, which may cause errors in fiber tracking. 2) Many brain tumors are remarkably heterogeneous [28, 11, 10, 27]. Particularly, the edema region surrounding the tumor may cause artefacts in fiber tracking, as the FA value is commonly affected in these regions [17].

### 1.3 Related work

The conventional tractography methods include Fiber Assignment by Continuous Tracking (FACT) and Unscented Kalman Filter (UKF) tractography.

#### FACT tractography

FACT [14] is a deterministic method that tracks fiber streamlines from a seed region by following the primary eigenvector from one voxel to the next. FACT is highly sensitive to the changes of FA and tensor values in the white matter. As a result, the tracking frequently fails when encountering peritumoral edema, leading to overestimated disruption of the structural networks. In some other cases, it may produce spurious tract rings around the tumor, which may underestimate the connectivity reduction.[6]

#### UKF tractography

UKF is shown to be able to track inside the peritumoral edema using two tensor models [12]. However, in order to achieve this, the model weakly controls the false positive fibers compared to healthy subjects [6]. The trade-off between false positive and false negative rates is challenging to be optimized among patients, which could particularly cause bias in the group analysis when individual tumors have heterogeneous extent of peritumoral edema.

### 1.4 Our contributions

We propose a method of estimating structural connectivity for network construction in brain tumor patients, without directly fiber tracking on patients (Fig. 1). Inspired by a method for traumatic brain injury [20], we firstly performed tratography on healthy controls and generated an unbiased tract template, with the location of each tract fiber derived. The TBSS approach was then employed to derive the skeletonized FA maps from patients, for estimating the connectivity strength of each specific tract in patients. To mitigate the bias in TBSS voxel projection caused by tumor mass effect, we further proposed an improved TBSS tailored to brain tumors, using an iterative projection of FA voxels guided by the tract orientation based on the voxel-wise eigenvector.

**Fig. 1.**
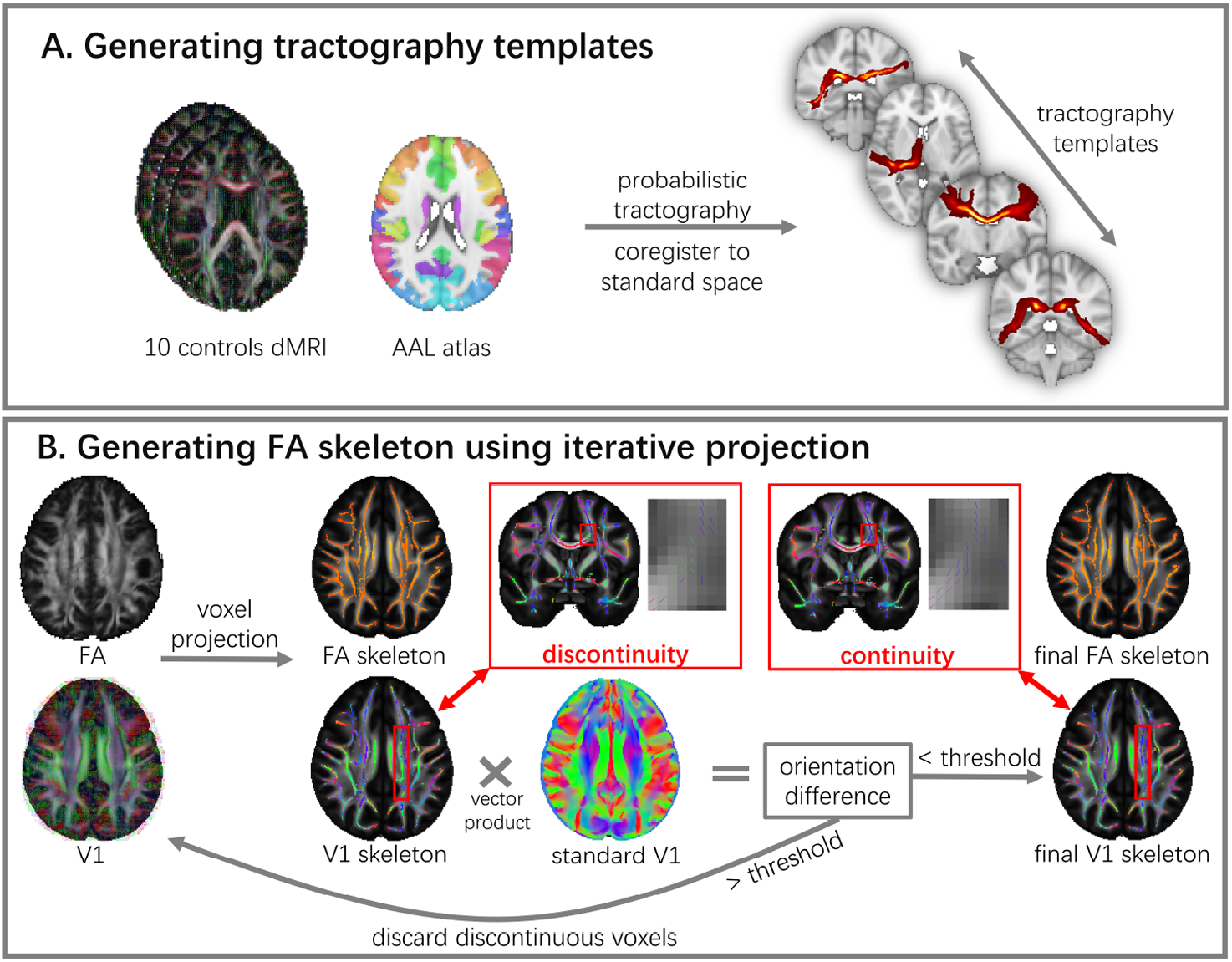
Flowchart of network construction. A. An unbiased tractography template is generated using probabilistic tractography in ten healthy subjects to indicate the location of the tracts between brain regions parcellated using the brain atlas. B. By using an iterative projection approach that considers the direction of voxles (V1), a skeletonized FA map with maximum tract continuity can be produced. The red arrows indicate the discontinuity in the FA skeleton from the traditonal TBSS that is corrected with our proposed method

We compared the connectivity strength estimated by the proposed approach and the conventional tractography methods in multiple datasets of brain tumor patients. The results show that our approach is reflecting more functional relevance. Further, the different extent of network disruption revealed by our approach correspond to the clinical prior knowledge of tumor histology.

## 2 Methods

### 2.1 Datasets

Three datasets of glioma or meningioma patients were included, with both diffusion MRI (dMRI) and resting-state functional MRI (rs-fMRI) available: 1) 4 low-grade gliomas (LGGs), 5 high-grade gliomas (HGGs) and 2 meningiomas (dMRI: 60 directions, b = 1000 s/mm2; rs-fMRI : TR = 2.5 s) were obtained from [16]; 2) OpenNeuro database: 7 LGGs, 4 HGGs, and 14 meningiomas (dMRI: 101 directions, b = 0, 700, 1200, 2800 s/mm2; rs-fMRI : TR = 2.4 s) [2]. 3) an in-house dataset: 12 HGGs (dMRI: 12 directions for each b values, b = 0, 350, 650, 1000, 1300, 1600 s/mm2; rs-fMRI : TR = 2.43 s). In total, 11 LGGs, 21 HGGs and 16 meningiomas were included.

### 2.2 Template-based brain networks

The proposed method includes three main steps:

– Producing an unbiased tractography template (Fig. 1A),
– Generating individualized FA skeleton (Fig. 1B),
– Combining tractography template with FA skeleton to estimate the connectivity strength between each pair of brain regions in patients

#### Tractography template

An unbiased tract template was generated in four steps using ten age-matched elderly healthy controls obtained from the Alzheimer’s disease Neuroimaging Initiative (ADNI, http://adni.loni.usc.edu/).

1. A diffusion model was fitted at each voxel of dMRI using Bedpostx of FM-RIB software library (FSL, https://fsl.fmrib.ox.ac.uk/fsl/). Pairwise probabilistic tractography was performed between 90 regions of Automated Anatomical Labelling atlas (AAL) atlas [22] using Probtrackx2 to generate a path distribution map in healthy controls [5].
2. The path distribution map was nonlinearly tranformed to standard MNI-152 space [7] using the Advanced Normalization Tools (ANTs)[3] and finally averaged, to produce a group distribution map for each tract. The maps were thresholded and binarized to preserve the top 5% strongest connections and generate a conservative tract atlas in the standard space.

#### Individual FA skeleton from iterative projection

An FA skeleton of each patient was generated at the individualized native space using an improved TBSS approach.

1. FA maps of all patients were first non-linearly co-registered to the standard space using ANTs. In addition, the corresponding principal eigenvector V1 maps were also transformed to the standard space. Instead of using the patient group thinned FA skeleton, we used the standard FA skeleton of FSL as the target for projection to reduce bias introduced by tumor.
2. We further improved TBSS that only projects the voxels with local maximum value at the tract center on the FA skeleton. Specifically, we compared the principal eigenvector (V1) of the projected voxel to the the standard V1 and calculated the orientation difference using vector product. The voxels with the orientation difference over 90 degrees were discarded, where neighbor voxels were selected instead. The projection iteration continued until all voxels on the skeleton converged to minimum direction difference with the standard FA skeleton. Using this procedure, the continuity of the voxel directions on each tract could be improved. An individualized FA skeleton was generated in each patient.

#### Structural connectivity strength estimation

The segments of the individualized FA skeleton within each tract of the template was extracted and averaged, representing the connectivity strengths of the major tracts.

### 2.3 Baseline methods

FACT was performed using diffusion toolkits [25], while UKF was performed using the UKFTratography packages in 3D Slicer(https://www.slicer.org/) via the SlicerDMRI project (http://dmri.slicer.org) [30, 15]. For both methods, the mean FA value of the tracts connecting two ROIs of AAL atlas was calculated using MRTrix3 by sampling and averaging the FA values along the streamlines generated by the tractography [21].

### 2.4 Functional brain networks

We constructed functional networks from rs-fMRI using GRETNA[23]. Firstly, the rs-fMRI signals were regressed, bandpass-filtered, smoothed with a kernel with a full width at half maximum of 6mm and co-registered to the standard space. Secondly, the brain regions of each patient were parcellated using the AAL atlas. Finally, pairwise Pearson correlation was performed between the mean rs-fMRI signals in 90 brain regions.

### 2.5 Functional relevance of the structural networks

#### Whole-brain structural and functional connectivity coupling

To measure the individualized functional relevance of a structural network, we calculated the whole brain structural connectivity-functional connectivity (SC-FC) coupling. Spearman rank correlation was performed between the non-zero elements of the structural connections with their corresponding functional connections to produce correlation coefficient for each patient. To compare the group difference, the correlation coefficients were transformed using Fisher Z-Transformation.

#### Group-wise structural-functional connection correlation

The strength of functional connections varies due to the distinct extent of disruptions in corresponding structural connections. By testing the correlation of each functional and structural connection, the functional connections sensitive to structural damage can be identified. To weakly control the family-wise error in multiple comparisons, we used the network-based statistics (NBS)[29], providing higher sensitivity in controlling false discovery rate.

### 2.6 Clinical validation

The clinical prior knowledge establishes that meningiomas normally cause less disturbance to the brain than gliomas. A robust network construction method should be sensitive to the difference between meningioma and glioma. We used the global efficiency, calculated according to graph theory, to characterize the brain networks, which were compared in meningioma and glioma patients [9].

## 3 Results

### 3.1 Iterative TBSS projection

The proposed approach showed smaller orientation differences between patient FA skeleton and standard FA, compared to the baseline methods (Table 1). One example is illustrated on Fig. 2. Tracts that are displaced by the tumor have voxels from the wrong directions projected onto the FA skeleton. In comparison, our approach could ensure the maximum orientation continuity of the tracts.

**Table 1.** Mean orientation differences between patient FA skeleton and standard FA

| Traditional TBSS projection<br>(radians) | Iterative TBSS projection<br>(radians) | *P*-value |
| --- | --- | --- |
| $0.424 \pm 0.057$ | $0.388 \pm 0.035$ | $3.8e-4$ |

**Fig. 2.**
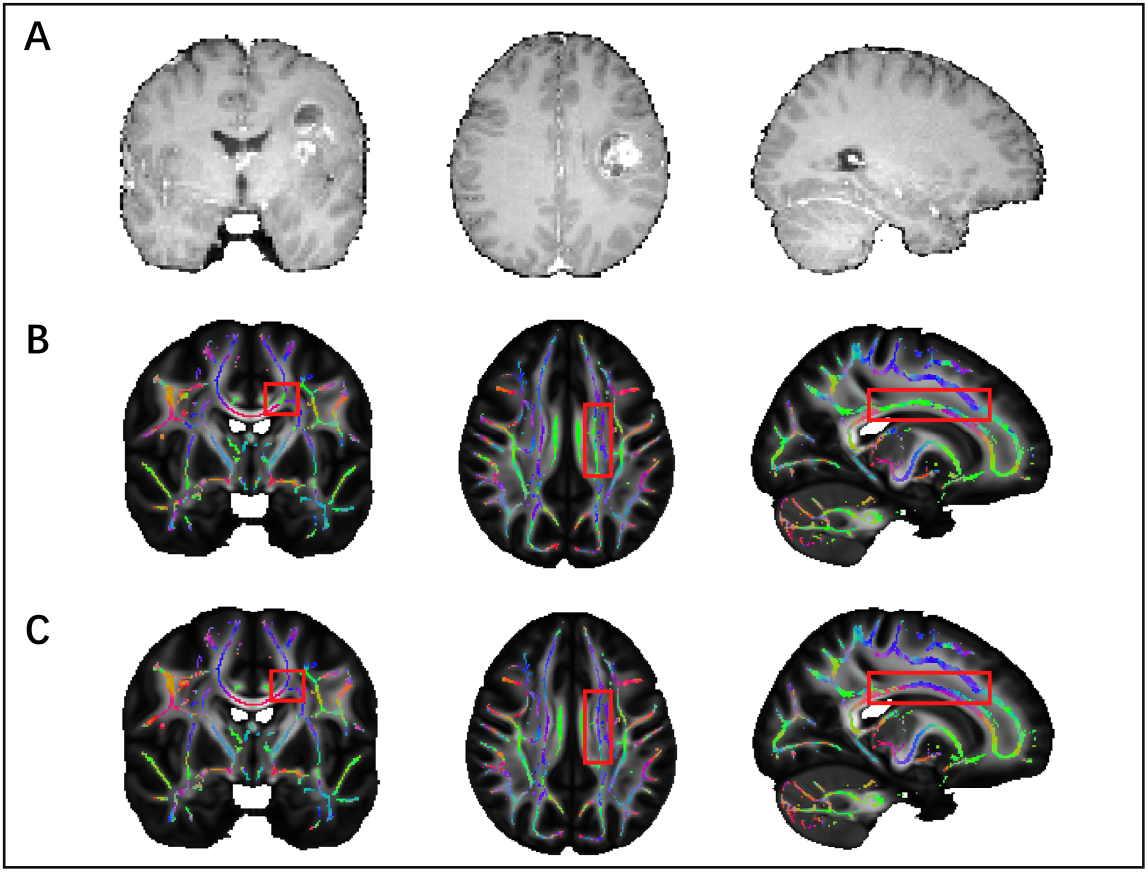
Example of voxel projection in coronal (left), axial (middle) and sagittal (right) views. A. T1 contrast MRI indicating tumor location. B. Traditional TBSS voxel projection: the tracts surrounding the tumor display different direction (green) from the contralesional tract (blue). C. The proposed method improves the direction continuity of the tract.

### 3.2 Whole brain structural and functional connectivity coupling

The proposed method achieved higher SC-FC coupling over the baseline methods (Table 2), suggesting the robustness of the brain network constructed using the proposed method.

**Table 2.** SC-FC Coupling coefficient (z-transformed)

| Methods | Mean ± SD | *P*-value (vs FACT) | *P*-value (vs UKF) |
| --- | --- | --- | --- |
| Proposed | $0.25 \pm 0.07$ | $3.3e-26$ | $3.5e-8$ |
| UKF | $0.16 \pm 0.08$ | $3.8e-10$ | - |
| FACT | $0.07 \pm 0.05$ | - | - |

### 3.3 Group-wise structural-functional connection correlation

Only the proposed method identified significantly correlated functional and structural sub-networks across the patient group, suggesting its high sensitivity to functional related structural disruptions.(Fig.3)

**Fig. 3.**
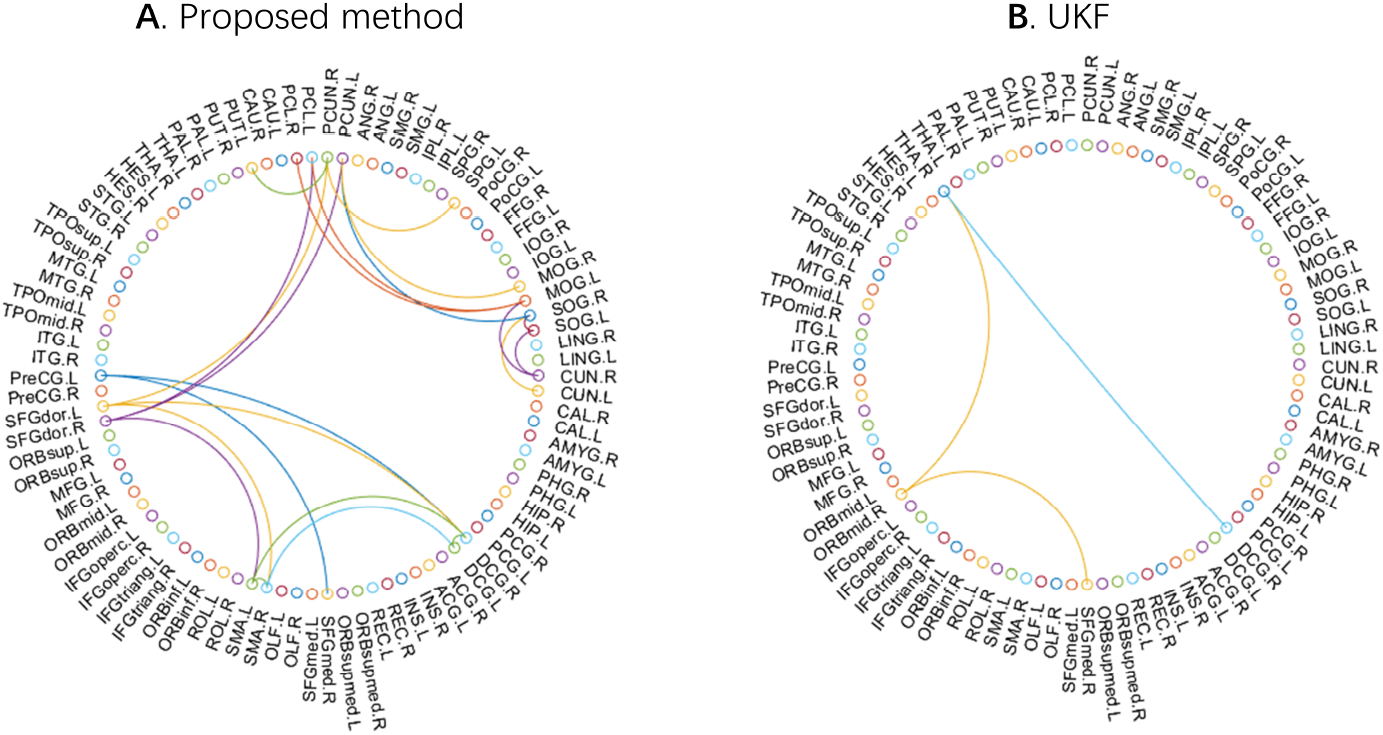
A. Sub-network is identified by the proposed method, with clusters of significantly correlated structural and functional connections (*P* = 0.0014). B. Fewer correlated structural and functional connections are identified by the UKF (*P >* 0.05). Family wise errors of both methods are corrected by NBS with 5000 permuations.

### 3.4 Global efficiency of different tumor types

The meningioma group in general has significantly higher global efficiency than the glioma group (*P* = 2.9e-4), while the baseline methods did not capture significant difference in Fig. 4. Further, in a subgroup of menigioma and high-grade glioma with comparable size, menigioma patients still have significant higher global efficiency comparing to high-grade glioma group (*P* = 9.3e-5), while baseline methods did not show significant difference.

**Fig. 4.**
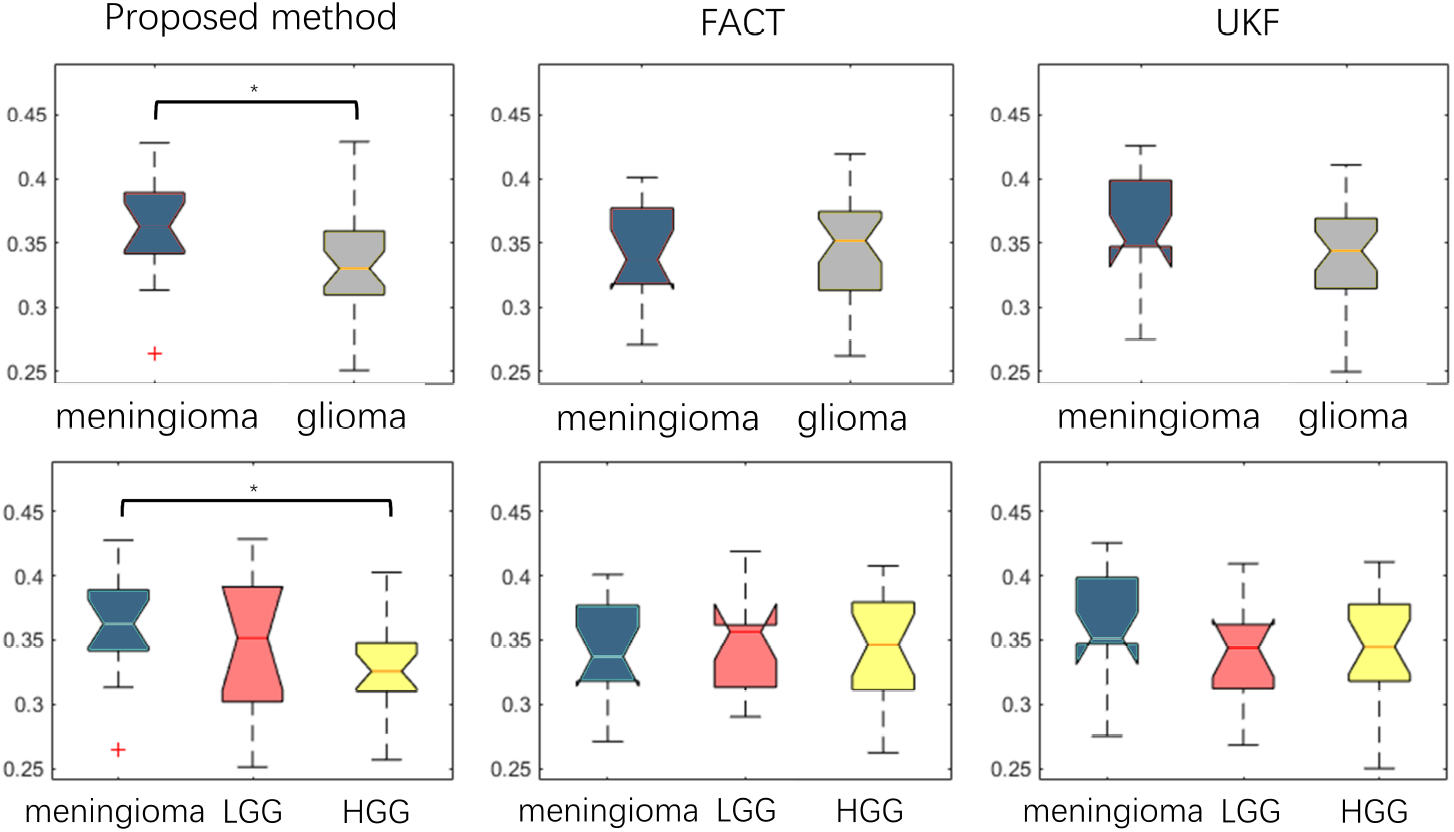
Global efficiency of the structural networks constructed using the proposed method, FACT, and UKF. Compared to glioma, meningioma patients display significantly higher global efficiency in the network constructed using the proposed approach.* : *P <* 0.05

## 4 Conclusion

This study proposes an approach to construct structural network and estimate the connectivity, which employs an improved TBSS approach and the tractography template from healthy controls. This method is shown to be robust compared to the conventional tractography, with higher functional and clinical relevance. The proposed method shows promise to aid treatment planning and patient risk assessment.

## Data Availability

The datasets generated during and/or analysed during the current study are available from the corresponding author on reasonable request

## Notes

### Competing Interest Statement

The authors have declared no competing interest.

### Funding Statement

SJP acknowledges National Institute for Health Research (NIHR) Career Development Fellowship (CDF-18-11-ST2-003). CL acknowledges Cancer Research UK biomarker grant (CRUK/A19732).

### Author Declarations

Ethical Committee: The Cambridgeshire 2 LREC, Ethics number: 10/H308/23

## References

1. Aerts, H., Fias, W., Caeyenberghs, K., Marinazzo, D.: Brain networks under attack: robustness properties and the impact of lesions. Brain 139 (12), 3063–3083 (2016)

2. Aerts, H., Marinazzo, D.: Brain tumor connectomics data (2019). https://doi.org/10.18112/openneuro.ds001226.v1.0.0

3. Avants, B.B., Tustison, N., Song, G.: Advanced normalization tools (ants). Insight j 2 (365), 1–35 (2009)

4. Bach, M., Laun, F.B., Leemans, A., Tax, C.M., Biessels, G.J., Stieltjes, B., Maier-Hein, K.H.: Methodological considerations on tract-based spatial statistics (tbss). Neuroimage 100, 358–369 (2014)

5. Behrens, T.E., Berg, H.J., Jbabdi, S., Rushworth, M.F., Woolrich, M.W.: Probabilistic diffusion tractography with multiple fibre orientations: What can we gain? Neuroimage 34 (1), 144–155 (2007)

6. Chen, Z., Tie, Y., Olubiyi, O., Rigolo, L., Mehrtash, A., Norton, I., Pasternak, O., Rathi, Y., Golby, A.J., O’Donnell, L.J.: Reconstruction of the arcuate fasciculus for surgical planning in the setting of peritumoral edema using two-tensor unscented kalman filter tractography. NeuroImage: Clinical 7, 815–822 (2015)

7. Grabner, G., Janke, A.L., Budge, M.M., Smith, D., Pruessner, J., Collins, D.L.: Symmetric atlasing and model based segmentation: an application to the hippocampus in older adults. In: International Conference on Medical Image Computing and Computer-Assisted Intervention. pp. 58–66. Springer (2006)

8. Incekara, F., Satoer, D., Visch-Brink, E., Vincent, A., Smits, M.: Changes in language white matter tract microarchitecture associated with cognitive deficits in patients with presumed low-grade glioma. Journal of neurosurgery 130 (5), 1538– 1546 (2018)

9. Latora, V., Marchiori, M.: Efficient behavior of small-world networks. Physical review letters 87 (19), 198701 (2001)

10. Li, C., Wang, S., Yan, J.L., Piper, R.J., Liu, H., Torheim, T., Kim, H., Zou, J., Boonzaier, N.R., Sinha, R., et al.: Intratumoral heterogeneity of glioblastoma infiltration revealed by joint histogram analysis of diffusion tensor imaging. Neurosurgery 85 (4), 524–534 (2019)

11. Li, C., Wang, S., Yan, J.L., Torheim, T., Boonzaier, N.R., Sinha, R., Matys, T., Markowetz, F., Price, S.J.: Characterizing tumor invasiveness of glioblastoma using multiparametric magnetic resonance imaging. Journal of neurosurgery 132 (5), 1465–1472 (2019)

12. Liao, R., Ning, L., Chen, Z., Rigolo, L., Gong, S., Pasternak, O., Golby, A.J., Rathi, Y., O’Donnell, L.J.: Performance of unscented kalman filter tractography in edema: Analysis of the two-tensor model. NeuroImage: Clinical 15, 819–831 (2017)

13. Liu, Y., Yang, K., Hu, X., Xiao, C., Rao, J., Li, Z., Liu, D., Zou, Y., Chen, J., Liu, H.: Altered rich-club organization and regional topology are associated with cognitive decline in patients with frontal and temporal gliomas. Frontiers in human neuroscience 14, 23 (2020)

14. Mori, S., Crain, B.J., Chacko, V.P., Van Zijl, P.C.: Three-dimensional tracking of axonal projections in the brain by magnetic resonance imaging. Annals of Neurology: Official Journal of the American Neurological Association and the Child Neurology Society 45 (2), 265–269 (1999)

15. Norton, I., Essayed, W.I., Zhang, F., Pujol, S., Yarmarkovich, A., Golby, A.J., Kindlmann, G., Wassermann, D., Estepar, R.S.J., Rathi, Y., et al.: Slicerdmri: open source diffusion mri software for brain cancer research. Cancer research 77 (21), e101–e103 (2017)

16. Pernet, C.R., Gorgolewski, K.J., Job, D., Rodriguez, D., Whittle, I., Wardlaw, J.: A structural and functional magnetic resonance imaging dataset of brain tumour patients. Scientific data 3 (1), 1–6 (2016)

17. Schult, T., Hauser, T.K., Klose, U., Hurth, H., Ehricke, H.H.: Fiber visualization for preoperative glioma assessment: Tractography versus local connectivity mapping. Plos one 14 (12), e0226153 (2019)

18. Sinha, R., Dijkshoorn, A.B., Li, C., Manly, T., Price, S.J.: Glioblastoma surgery related emotion recognition deficits are associated with right cerebral hemisphere tract changes. Brain communications 2 (2), fcaa169 (2020)

19. Smith, S.M., Jenkinson, M., Johansen-Berg, H., Rueckert, D., Nichols, T.E., Mackay, C.E., Watkins, K.E., Ciccarelli, O., Cader, M.Z., Matthews, P.M., et al.: Tract-based spatial statistics: voxelwise analysis of multi-subject diffusion data. Neuroimage 31 (4), 1487–1505 (2006)

20. Squarcina, L., Bertoldo, A., Ham, T.E., Heckemann, R., Sharp, D.J.: A robust method for investigating thalamic white matter tracts after traumatic brain injury. Neuroimage 63 (2), 779–788 (2012)

21. Tournier, J.D., Smith, R., Raffelt, D., Tabbara, R., Dhollander, T., Pietsch, M., Christiaens, D., Jeurissen, B., Yeh, C.H., Connelly, A.: Mrtrix3: A fast, flexible and open software framework for medical image processing and visualisation. NeuroImage 202, 116137 (2019)

22. Tzourio-Mazoyer, N., Landeau, B., Papathanassiou, D., Crivello, F., Etard, O., Delcroix, N., Mazoyer, B., Joliot, M.: Automated anatomical labeling of activations in spm using a macroscopic anatomical parcellation of the mni mri single-subject brain. Neuroimage 15 (1), 273–289 (2002)

23. Wang, J., Wang, X., Xia, M., Liao, X., Evans, A., He, Y.: Gretna: a graph theoretical network analysis toolbox for imaging connectomics. Frontiers in human neuroscience 9, 386 (2015)

24. Wang, J., Xu, S.L., Duan, J.J., Yi, L., Guo, Y.F., Shi, Y., Li, L., Yang, Z.Y., Liao, X.M., Cai, J., et al.: Invasion of white matter tracts by glioma stem cells is regulated by a notch1–sox2 positive-feedback loop. Nature neuroscience 22 (1), 91–105 (2019)

25. Wang, R., Benner, T., Sorensen, A.G., Wedeen, V.J.: Diffusion toolkit: a software package for diffusion imaging data processing and tractography. In: Proc Intl Soc Mag Reson Med. vol. 15. Berlin (2007)

26. Wei, Y., Li, C., Cui, Z., Mayrand, R.C., Zou, J., Wong, A.L., Sinha, R., Matys, T., Schönlieb, C.B., Price, S.J.: Structural connectome quantifies tumor invasion and predicts survival in glioblastoma patients. bioRxiv (2021)

27. Yan, J.L., Li, C., Boonzaier, N.R., Fountain, D.M., Larkin, T.J., Matys, T., van der Hoorn, A., Price, S.J.: Multimodal mri characteristics of the glioblastoma infiltration beyond contrast enhancement. Therapeutic advances in neurological disorders 12, 1756286419844664 (2019)

28. Yan, J.L., Li, C., van der Hoorn, A., Boonzaier, N.R., Matys, T., Price, S.J.: A neural network approach to identify the peritumoral invasive areas in glioblastoma patients by using mr radiomics. Scientific reports 10 (1), 1–10 (2020)

29. Zalesky, A., Fornito, A., Bullmore, E.T.: Network-based statistic: identifying differences in brain networks. Neuroimage 53(4), 1197–1207 (2010)

30. Zhang, F., Noh, T., Juvekar, P., Frisken, S.F., Rigolo, L., Norton, I., Kapur, T., Pujol, S., Wells III, W., Yarmarkovich, A., et al.: Slicerdmri: Diffusion mri and tractography research software for brain cancer surgery planning and visualization. JCO clinical cancer informatics 4, 299–309 (2020)

